# Anatomy of a Failure: A Retrospective Evaluation of a Cognitive Bias Modification Intervention to Promote Physical Activity in Cardiac Rehabilitation

**DOI:** 10.1101/2025.02.27.25323019

**Authors:** Layan Fessler, Silvio Maltagliati, Philippe Meyer, Axel Finckh, Stéphane Cullati, David Sander, Malte Friese, Reinout W. Wiers, Ata Farajzadeh, Christophe Luthy, Philippe Sarrazin, Boris Cheval

## Abstract

**Objectives:** Promoting regular physical activity (PA) is essential in cardiac rehabilitation; yet many patients exhibit low levels of PA. In January 2022, the Improving Physical Activity (IMPACT) trial, a randomised controlled trial at the University Hospital of Geneva, was launched to promote PA in cardiac patients by targeting automatic approach tendencies towards exercise-related stimuli through a cognitive bias modification (CBM) intervention. This article examines the limited acceptance of this intervention, identifies potential barriers, and proposes strategies to improve future implementations.

**Design:** Retrospective evaluation of a pre-registered clinical trial.

**Setting:** The intervention was conducted in a cardiac rehabilitation centre in Switzerland. **Participants.** Sixty-eight cardiac rehabilitation patients (*M*_age_ = 57.76, SD = 10.76 years, 87% male).

**Intervention:** Patients received 12 CBM sessions over 6 weeks, designed to target approach-avoidance tendencies to exercise-related stimuli and improve PA levels.

**Primary and secondary outcome measures:** Acceptance was assessed using behavioural (e.g., enrolment and engagement rates), cognitive (e.g., perceived effectiveness), and emotional (e.g., affective evaluation) indicators. The cognitive and emotional indicators were derived from verbal feedback documented by the research assistants based on patients’ reactions during the intervention period. These observations do not constitute qualitative research as defined by methodological standards; they were informal notes provided by RAs during intervention delivery and were not collected or analysed using qualitative research methods.

**Results:** Of the 352 patients initially required, only 68 (19%) were enrolled. Among these 68, 63% completed the minimum number of CBM sessions, and 25% completed accelerometer-based PA measures during the week following discharge. These behavioural indicators of low acceptance showed cognitive (e.g., scepticism about the task relevance and perceived effectiveness of the intervention) and emotional (e.g., feelings of boredom and disinterest) barriers.

**Conclusion:** The low engagement and acceptance observed in the IMPACT trial reveal highlight several key barriers, such as perceived task irrelevance, task monotony, and task boredom, that undermine the acceptance and feasibility of this digital CBM intervention in cardiac rehabilitation. These findings emphasise the importance of designing patient-centred interventions, ensuring their seamless integration into clinical contexts, and conducting qualitative research prior to implementation to anticipate potential barriers.

**Strengths and limitations of this study:**

- This study uses a multidimensional assessment of acceptance by examining behavioural, cognitive, and emotional indicators of a cognitive-bias modification intervention.
- Including informal verbal feedback from research assistants (RAs) offer valuable insights into patients’ experience and perception of the intervention in a real-world cardiac rehabilitation setting.
- Acceptance was assessed primarily through RAs’ verbal feedback collected during intervention delivery, rather than direct patients’ reports or standardised measures, which may introduce recall bias and limit the reliability of the findings.

## Introduction

Promoting physical activity (PA) in patients with cardiovascular diseases is critical, as regular PA is associated with significant health benefits, including reduced risks of mortality, less frequent cardiac events and hospitalisations, and improved quality of life.^1–3^ Despite these benefits, many patients exhibit low levels of PA, and previous interventions have failed to produce sustained behavioural changes.^4–8^ Therefore, it is essential to design, implement, and evaluate innovative interventions aimed at promoting PA behaviour in this population. Along with more reflective factors (e.g., intention to be physically active), recent research highlights the potential of targeting automatic precursors of PA, such as automatic action tendencies, to enhance interventions effectiveness.^9^ Automatic tendencies, grounded in dual-process accounts of human behaviour,^10–12^ refer to the automatic preparation of behavioural schemata, directing the organism’s action towards or away from an object.^13^ While extensively studied in psychopathology, – where approach tendencies are linked to addictions and avoidance tendencies to anxiety disorders^14^ –, emerging evidence suggests that they also play an important role in health behaviours, including PA,^15–20^ as they influence individuals’ inclination to engage in or to avoid effortful activities. Interventions based on Cognitive Bias Modification (CBM) may be particularly beneficial for patients recovering from an acute health event, such as a stroke or heart attack. This population is often more susceptible to fear of PA,^21^ which can result in avoidance behaviours.^22^ However, to our knowledge, no randomised controlled trials (RCTs) have been conducted to evaluate the efficacy of such interventions in patients with cardiovascular diseases. The Improving Physical Activity (IMPACT) trial was designed to address this critical gap in the literature.^23^

The main purpose of this article is to report on the acceptance of the IMPACT trial, which was based on a CBM intervention for cardiac rehabilitation patients. We then propose strategies to improve effectiveness of future interventions.

## Methods

### The IMPACT Trial

The protocol of the IMPACT trial, including its theoretical background and planned analyses, has been detailed elsewhere.^23^ In brief, the IMPACT trial was designed to target approach-avoidance tendencies towards exercise-related stimuli using a CBM intervention for patients enrolled in a cardiac rehabilitation programme. Similar CBM interventions have shown promise in reducing certain unhealthy addictive behaviours such as alcohol consumption and smoking;^24–27^ and in promoting healthy behaviours such as healthy eating and PA.^26,28–31^

The IMPACT trial was conducted at the University Hospital of Geneva in Switzerland. Ethical approval was granted by the Geneva Cantonal Ethics Committee (reference CCER2019-02257). This approval covers the present study, including the collection and analysis of additional data. Specifically, these data include a retrospective, team-level reflection on implementation barriers encountered during the intervention. These reflections are based on aggregated, non-systematic observations and verbal feedback from patients and staff, with no individual-level data collected or stored.

All participants provided written informed consent to participate in the IMPACT trial and for their data to be used in future research. Patients were eligible for inclusion if they were: (a) receiving treatment in an outpatient cardiac rehabilitation programme; (b) aged 18 years or older; (c) were physically and cognitively able to complete the study protocol; and (d) provided written informed consent to participate in the IMPACT trial and for their data to be used in future research. Patients were excluded if they had a medical contraindication to physical activity. All eligibility criteria were verified by the medical team. The intervention initially consisted of a training programme of 12 sessions over 3 weeks (i.e., an average of 4 sessions per week) using a Visual Approach/Avoidance by the Self Task (VAAST^32^).

However, logistical constraints – particularly a limited number of research assistants – restricted the participant pool to ambulatory patients and extended the programme duration to 6 weeks. Consequently, the intervention was adapted to 12 sessions over 6 weeks (i.e., 2 sessions per week). In the VAAST, patients responded to the format (i.e., portrait vs landscape format) of images depicting PA or sedentary behaviours by pressing a button on a touchscreen tablet to virtually move forward or backward in the image, thereby approaching or avoiding the stimuli. Patients were instructed to approach the image when it appeared in a portrait format and to avoid the image when it appeared in a landscape format, with the rule being counterbalanced across participants. Participants completed the VAAST in the presence of research assistants, not privately in front of a private computer over the Internet.

In the intervention group, patients received training in which 90% of images depicting PA were presented in the approach format (and 10% in the avoidance format), while 90% of images depicting sedentary behaviours were presented in the avoidance format (and 10% in the approach format). In contrast, the comparator group (placebo; sham controlled) approached and avoided PA and sedentary behaviours equally often (50% in the approach format and 50% in the avoidance format), so they were neither specifically trained to approach PA nor to avoid sedentary behaviours, compared to patients in the intervention group. At the end of the 6 weeks of rehabilitation, patients were asked to wear an accelerometer over 1 week to measure time spent in moderate-to-vigorous PA, which served as the primary outcome of the IMPACT trial. The main hypothesis was that the CBM intervention group would show higher levels of PA during the week following rehabilitation discharge compared to the placebo intervention group.

### The Acknowledgement of the Failure

The IMPACT trial faced significant challenges, particularly regarding the acceptance of the intervention among patients, which is defined as a posteriori pragmatic evaluation of an activity (e.g., an intervention).^33^ As a low acceptance significantly hinders the success of an intervention, we deemed important to better understand the reasons underlying the low acceptance of our intervention. Given these challenges, the IMPACT trial has since been terminated as it has proven unfeasible in its original form. In this context, the Theoretical Framework of Acceptability^a^ (TFA^34^) posits that acceptance can be assessed through behavioural indicators (e.g., attrition), cognitive indicators (e.g., scepticism about the task relevance and perceived effectiveness of the intervention), and emotional indicators (e.g., feelings of boredom, disinterest, or frustration). It should be noted that the TFA uses the term “acceptability” to refer to the evaluation of an intervention before, during, and after its implementation.^34^ However, some authors propose a distinction between the a priori evaluation of a task (acceptability) and the actual use of the task (acceptance).^33^ In this report we have used only the term “acceptance” to ensure internal consistency and readability. This choice was made because patient feedback was primarily reported during the intervention period, not before. Understanding and addressing these behavioural, cognitive, and emotional dimensions of acceptance is critical to improving the design and implementation of interventions. Therefore, the main objective of the present article is to report on the low acceptance of the IMPACT trial and the CBM intervention in cardiac rehabilitation patients, investigate potential reasons for these challenges, and propose strategies to improve the potential of future interventions.

### Patient and public involvement

Patients and/or the public were not involved in the design, conduct, reporting or dissemination of this study.

### Measures

In this Methods section, we describe only the behavioural, cognitive and emotional indicators of acceptance of the IMPACT trial. The methodology used to assess demographic, anthropometric and psychological outcomes is described elsewhere.^35^ It is important to note that the objective of this report was defined after the trial was halted due to insufficient participant engagement. As a result, patients’ verbal feedback was not included in the initial protocol. (see Cheval et al.^23^). Feedback reports were compiled by research assistants during the intervention based on verbal reports from patients and medical staff, which were not recorded or documented at the time. These reports were not formally recorded or documented, nor were they obtained through structured methods such as questionnaires, surveys, or direct recordings. Instead, they consisted of unsystematic observations, including patients’ reactions, views of clinical staff, practical difficulties with the protocol and implementation barriers. These were verbally relayed by research assistants immediately after relevant interactions. Due to their informal and unstructured nature, these observations do not adhere to the methodological standards of qualitative research. Instead, they served as contextual documentation that emerged organically during the intervention’s delivery. We analyse them here as supplementary contextual information to better understand implementation barriers, while explicitly acknowledging their methodological limitations. This reliance on research assistants’ recollections of patients’ verbal feedback may introduce recall bias, thereby limiting the validity and accuracy of these indicators.

### Behavioural Indicators

#### Non-Enrolment Rate

The non-enrolment rate was defined as the percentage of patients who attended the initial meeting with the research assistants and provided consent to participate in the IMPACT trial.

#### CBM Training Engagement Rate

The engagement rate in the CBM intervention was defined as the percentage of enrolled patients in the IMPACT trial (i.e., who agreed to participate in the IMPACT trial) who completed the minimum number of training sessions (i.e., 3) over the 6 weeks.

#### Engagement Rate in Post-Intervention PA Measures

PA engagement was defined as the percentage of patients with sufficient engagement to CBM training who provided complete PA measures in the week following the rehabilitation programme. PA was measured using an accelerometer (ActiGraph GT3X, Pensacola FL, USA) worn on the hip. PA measures were considered valid if the device was worn for at least 10 waking hours per day for at least four days, including one weekend day.^36,37^

For each of the 3 indicators, we compared the sample size achieved with the planned sample size of 352 to test our primary hypothesis. This was done to estimate the difference between the planned and actual sample sizes. For all these indicators, results were also described separately by group (intervention and placebo).

### Cognitive Indicators

#### Perceived Effectiveness of the Intervention

Prior to the start of the intervention, perceived effectiveness of the intervention was assessed using the following item: “To what extent do you think that your PA behaviours will improve as a result of training on the computerised task?”^23^ Participants answered on a scale from 1 (not at all) to 5 (totally). During the intervention period, research assistants documented any spontaneous patients’ and medical staff feedback related to perceived effectiveness of the intervention, with particular attention to verbal evaluations of the approach-avoidance training task.

#### Burden

Prior and during the intervention period, the burden of the intervention was assessed using patients’ and medical staff spontaneous feedback. Research assistants verbally reported any such feedback relating to the perceived effort required to participate in the intervention to the research team, with particular attention to verbal evaluations of the task perceived difficulty or demands. However, this feedback was not (formally) recorded or documented by the research assistants.

### Emotional Indicators

#### Attitudes Towards the Intervention

Prior and during the intervention period, affective attitudes towards the intervention (i.e., the extent to which participants perceived the intervention as something unpleasant or pleasant, boring or fun) were assessed using patients’ spontaneous feedback. Research assistants verbally reported any spontaneous patient feedback related to their feelings about the intervention to the research team, with particular attention to verbal ratings of the level of boredom-fun and disinterest-interest towards the CBM task. This feedback was not (formally) recorded or documented by the research assistants.

### Resources Constraints

Resource limitations, such as insufficient equipment and a shortage of research assistants, were documented as potential barriers to the effective implementation of the intervention.

## Results

Descriptive characteristics of the sample are presented in Supplementary Material 1. A total of 68 outpatients enrolled in a cardiac rehabilitation programme were included in this evaluation. The majority were male (86.76%), with a mean age of 57.76 years (SD = 10.76). Mean left ventricular ejection fraction was 52.87% (SD = 10.49). The most common indication for cardiac rehabilitation programme was acute coronary syndrome, comprising 40.30% with non-ST-elevation myocardial infarction (NSTEMI) and 26.87% with ST-segment elevation myocardial infarction (STEMI). Nearly half of the participants (45.45%) were classified as overweight, and the majority reported engaging in some PA in daily life prior to programme enrolment. Patients also reported low-to-moderate levels of anxiety (*M* = 1.79, SD = 0.90, scale: 1–5), depressive symptoms (*M* = 1.62, SD = 0.76, scale: 1–5), fatigue (*M* = 2.55, SD = 1.06, scale: 1–5), and pain intensity (*M* = 2.10, SD, 2.29, scale: 1–5) in their daily life. The medical team reported that around 10% of patients were accompanied by a family member during programme sessions.

Comparative analyses of the intervention and control groups of participants with complete PA data at the end of the programme (n = 17) are presented in Supplementary Material 2 (Tables S2–S3). As expected, given the small sample size and implementation challenges, no statistically significant differences were observed between the groups for any outcome (all p > 0.05). However, we did observe a statistically significant difference in global perceived physical health (M_intervention_ = 2.50, SD = 0.71 vs. M_control_ = 3.71, SD = 1.11, p = .015, scale: 1–5). However, these analyses should be interpreted with caution due to substantial attrition and the study’s limited statistical power.

### Behavioural Indicators

In terms of non-enrolment rate, over the first 18 months after the trial launch, 262 patients attended the initial meeting during which the intervention was described. Of them, 68 patients (26%) agreed to participate in the IMPACT protocol. Regarding engagement in the CBM training, 43 out of the 68 patients who started the protocol (63%) completed the minimum required three training sessions over six weeks. As for engagement in the accelerometer-based measures of PA, 17 of the 43 patients (40%) who completed sufficient CBM sessions provided valid accelerometers data. As a result, relative to the 262 participants screened for enrolment, 26% (n = 68) were enrolled, 16% (n = 43) completed the CBM training, and only 6% (n = 17) provided PA data during the week following discharge from the rehabilitation programme (see Figures 1 and 2)^b^. Overall, the results showed a high attrition rate across the entire protocol, with 75% of enrolled participants (i.e., 51/68) withdrawing from the study before the end of the protocol.

**Figure 1.**
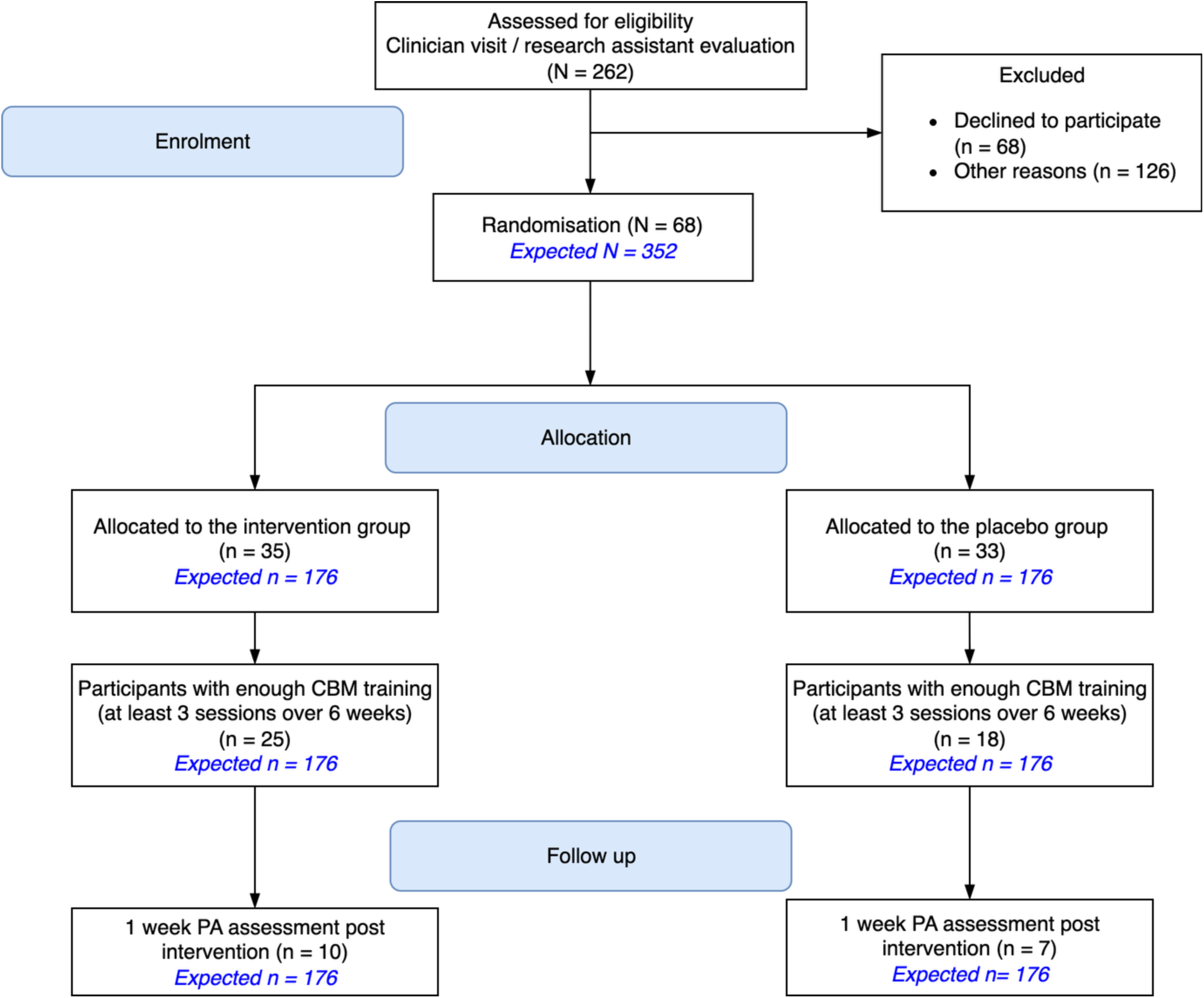
Flowchart of the Study. Abbreviations: CBM, cognitive bias modification; PA, physical activity. The expected sample size expected in Cheval et al.^23^ is shown in blue.

**Figure 2.**
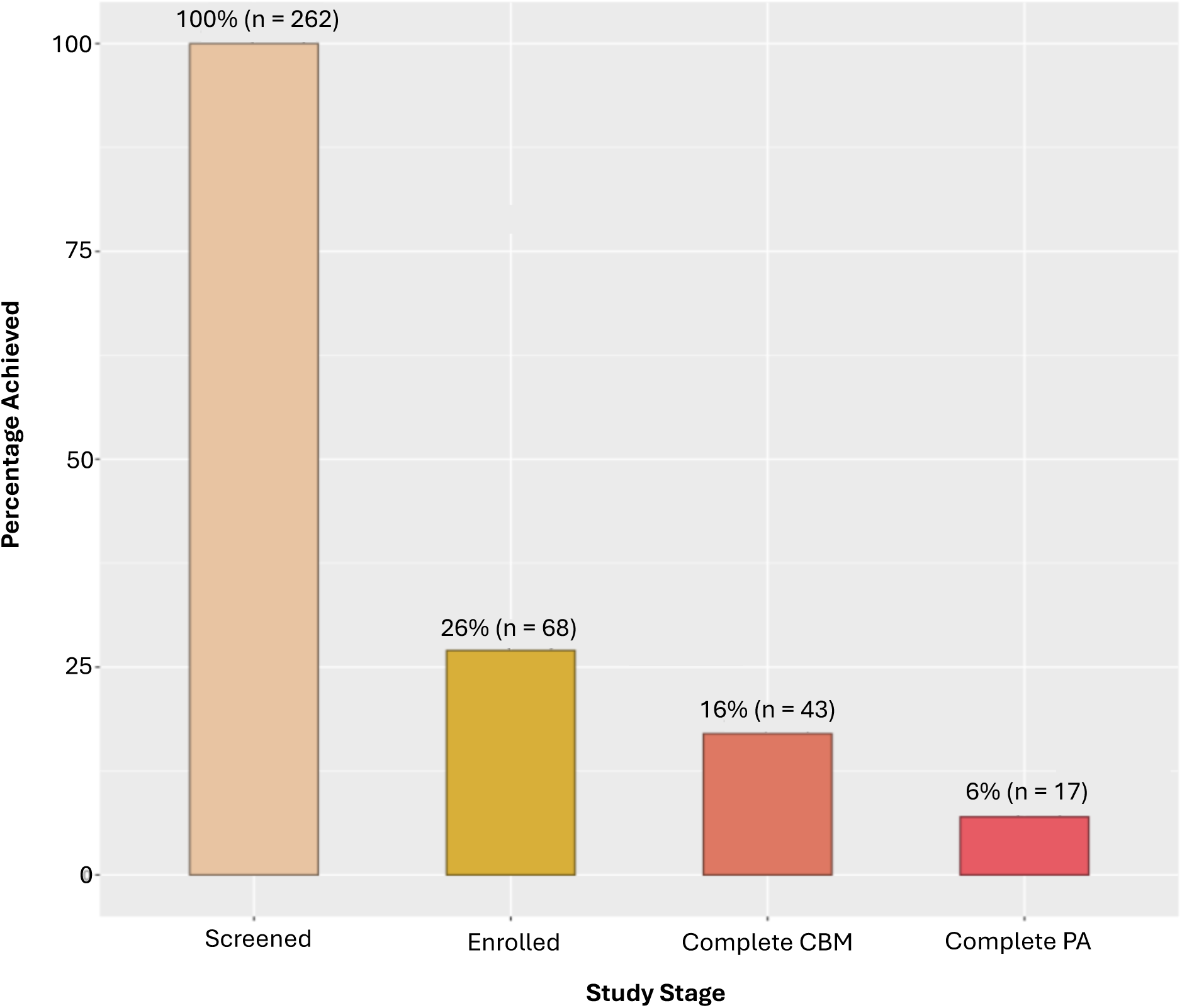
Study Progress Metrics. Abbreviations: Screened, patients who were screened and found eligible for the study; Enrolled, patients who were enrolled in the study; Complete CBM, participants who completed the minimum required CBM training (i.e., three sessions); Complete PA, participants who provided complete physical activity data during the week following discharge from the rehabilitation programme. Percentages were calculated on the screened patients.

Additional analyses were performed to examine the power achieved with a final sample of 17 patients (n = 10 patients in the intervention group and n = 7 in the placebo group). For a two-tailed t-test, an alpha of .05, an effect size of d = 0.35, and a total sample size of 17, we could expect a power of 10% (Supplementary Material 3, Figure S1).

We now present the results stratified by group (placebo and intervention). Thirty-five patients (51.5% of the total number of enrolled participants) were assigned to the intervention group. Regarding engagement in the CBM training, 25 out of the 35 patients who started the protocol (71%) completed the minimum required 3 training sessions over 6 weeks. When it came to engagement in the accelerometer-based measures of PA, only 10 of the 25 patients who completed the CBM training (40%) provided valid data. As a result, relative to the planned sample size of 176 patients per group, the intervention group reached 20% (n = 35) of the targeted enrolment, 14% (n = 25) for engagement in the CBM training, and a mere 6% (n = 10) for PA data in the week following discharge from the rehabilitation. Concurrently, 33 patients (48.5% of the total number of enrolled participants) were assigned to the placebo sham-controlled group. Regarding engagement in the CBM training, 18 out of the 33 patients who started the protocol (55%) completed the minimum required 3 training sessions over 6 weeks. When it came to engagement in the accelerometer-based measures of PA, 7 out of the 18 patients who completed the CBM training (39%) provided valid data. As a result, relative to the planned sample size of 176 patients per group, the placebo group only reached 19% (n = 33) of the targeted enrolment, 10% (n = 18) for engagement in the CBM training, and a mere 4% (n = 7) for collecting PA data in the week following discharge from the rehabilitation

### Cognitive Indicators

Prior to the start of the intervention period, patients demonstrated a moderate perceived effectiveness of the intervention (*M* = 3.09 on a scale ranging from 1 to 5; Supplementary Material 4, Table S4 and Figure S2). Informal feedback from research assistants indicated that some patients were sceptical about the effectiveness of the intervention. For instance, patients questioned how the task could help them become more active. Some were also reluctant to participate in activities they perceived as unnecessary, stating that they did not want to take on extra tasks during their rehabilitation. This reluctance was probably because of the demanding nature of the rehabilitation protocol, which was part of their standard clinical care (see Supplementary Material 5 for details).

Another factor that may have reinforced the perception of the intervention as an additional burden was the lack of integration of research assistants into the healthcare team. Despite our efforts to embed them within the clinical setting, research assistants were not fully integrated (e.g., attending weekly team seminars or receiving formal introductions as part of the clinical staff). This separation likely contributed to the intervention being viewed as an extra task, distinct from the regular rehabilitation protocol, thereby increasing the perceived burden. This separation likely increased the perceived burden not only for patients but also for healthcare professionals, who may have considered it an extra task outside their standard clinical responsibilities.

Additionally, informal feedback from research assistants suggests that medical staff, including doctors and physiotherapists, faced challenges in reminding patients to attend their scheduled CBM sessions. Staff reported that time constraints and competing priorities within the rehabilitation protocol hindered their ability to consistently prompt patients to participate in these sessions.

### Emotional Indicators

Additional feedback from research assistants highlighted significant issues related to patients’ boredom and disinterest. Patients reportedly found the CBM sessions monotonous and perceived them as long and unengaging. They also showed little interest in interacting with tablet-based activities. Furthermore, patients criticised the visual presentation of the CBM task, commenting that the images were unappealing and suggesting that a more colourful design might improve their experience. As the intervention progressed, boredom seemed to increase as feedback about monotony became more frequent. It is possible that the novelty of the intervention, a key factor in maintaining interest and curiosity,^38^ wore off over time, contributing to the increased feelings of boredom.

### Resources Constraints

A total of 8 tablets were available to deliver the CBM intervention to all patients, which created logistical challenges as the devices were occasionally in use when potential participants were available for recruitment. In addition, the 3 research assistants, who were students at the University of Geneva, were not available full-time, which limited their ability to recruit participants consistently.

## Discussion

The main purpose of this report was to examine the limited acceptance of the IMPACT trial, a CBM intervention in cardiac rehabilitation patients, to identify the underlying factors and, ultimately, to propose strategies to improve future interventions. We observed a low enrolment, with only 26% of the patients screened agreeing to participate, and low adherence, as 16% of participants completed the intervention and just 6% provided PA data. Key barriers to acceptance were identified, including cognitive factors, such as scepticism about the task’s relevance and the perceived effectiveness of the intervention, and emotional factors, such as feelings of boredom and disinterest. These findings suggest that such an intervention could be difficult to implement in a clinical context without significant changes. However, these results should be interpreted in the context of the informal, unplanned and unsystematic data collection process, which relied on the research assistants’ recollections of their interactions with patients.

### Difficulty with Patient Enrolment: Reasons and Recommendations for Future Studies

The first major challenge we faced was patient enrolment, a critical factor in the success of clinical trials.^34,39^ In clinical research, studies are often abandoned due to insufficient recruitment,^39^ and a clinicians survey identified it as the most significant difficulty in conducting clinical research.^40^ Our initial goal was to recruit a sample of 352 patients to adequately test our primary hypothesis. This number included 262 patients, with a buffer to account for a potential loss to follow-up of 10–20% 1-week after the end of the intervention. To maintain sufficient statistical power for the 1-year follow-up, we also anticipated a dropout rate of 30–40%, resulting in a planned sample size of 352. However, after 18 months, we were only able to recruit 68 patients, which was only 19% of the expected sample size and 26% of the patients screened for enrolment. This significant shortfall meant that, even before considering acceptance issues, we lacked the statistical power necessary to accurately test our hypothesis.

One possible cognitive indicator of this low enrolment and acceptance was the perceived burden of the intervention protocol. Patients may have seen it as a research initiative rather than an integral part of their rehabilitation programme, and may have perceived it as an additional constraint. This lack of seamless integration into their daily health care routines may have diminished its perceived relevance and importance, further discouraging participation and reducing their willingness to prioritise the CBM programme within their rehabilitation schedules.

Another cognitive indicator can be found in the reported moderate baseline intentions to engage in regular PA (M = 5.51, SD = 1.34 on a 0–10 scale; see Supplementary Table S1), which may have been insufficient to motivate engagement with an additional intervention specifically targeting PA behaviour. This modest motivation suggests that the programme may have been targeting patients who were not yet ready for such an intervention, or that the intervention itself failed to sufficiently enhance motivation. In addition, the lack of family involvement may have compounded these challenges further. Despite explicit invitations, the medical team reported that only 10% of patients were accompanied by a family member during programme sessions. Family involvement has been identified as a strong predictor of adherence to cardiac rehabilitation programmes,^41^ and its near absence in our study likely further undermined engagement. Finally, although cardiac rehabilitation is strongly recommended following a myocardial infarction in Switzerland, participation is voluntary rather than mandatory. Therefore, the combination of modest baseline motivation, limited external support from family members, and the voluntary nature of cardiac rehabilitation participation may have created an environment where an additional cognitive intervention, already perceived as burdensome rather than supportive, faced significant engagement barriers.

Moreover, some patients expressed scepticism about the intervention’s effectiveness and were discouraged by its digital format, particularly the use of touchscreen tablets. This perceived lack of relevance of the task probably contributed to the difficulties in recruiting sufficient participants for the IMPACT trial. For those who agreed to participate in the protocol, resource constraints further complicated recruitment. The limited availability of tablets – only 8 in total – often meant that devices were unavailable when patients were ready to engage. Additionally, the absence of research assistants in the department at all times hindered consistent communication and support, which may have further discouraged participation. Collectively, these indicators contributed to patients’ low acceptance of the intervention.

Recommendations for potential improvements in enrolment and acceptance are primarily based on the characteristics identified above (see Table 1 for a summary). To optimise patient enrolment, we suggest that future studies integrate the protocol as a central, albeit optional, component of the patient’s rehabilitation treatment.^42^ It is essential that healthcare professionals working with the patients, such as medical doctors, physiotherapist, and nurses actively engage in the study protocol. Future implementations should consider strategies to systematically involve family members in supporting patient engagement, assess baseline readiness for behaviour change interventions, and carefully consider how additional programmes are framed and integrated within existing rehabilitation structures.

**Table 1.**
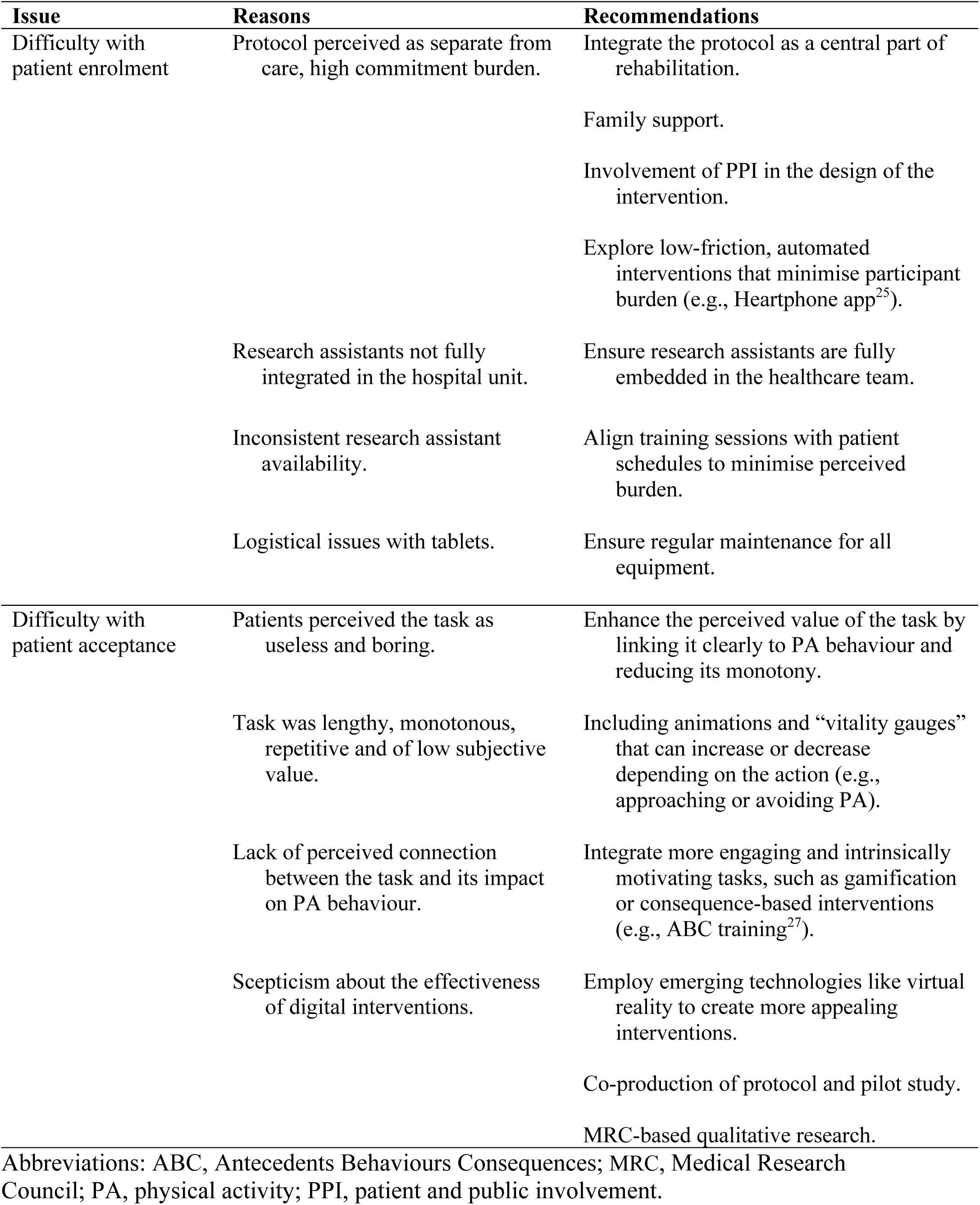
Potential Reasons for Failure and Recommendations for Future Studies.

Additionally, early and meaningful patient and public involvement (PPI) in the design and planning of interventions can provide critical insights into potential barriers and facilitators. PPI members help to ensure that interventions are patient-centred, feasible and acceptable by offering perspectives on the hospital environment, patient preferences and practical challenges that researchers might otherwise overlook.^43,44^ For instance, their input can inform the development of materials and the integration of the protocol into patients’ routines, reducing perceived burden and increasing engagement.

Research assistants should be fully integrated into the healthcare team and recognised as essential members by both the healthcare professionals and the patients. For example, ensuring that all training sessions are well integrated into patients’ schedules can help reduce the perception of additional burden. Notably, the protocol should be presented in such a way that patients feel that participation requires minimal effort and time.^34,42^

### Difficulty with Patient Acceptance: Reasons and Recommendations for Future Studies

The second challenge encountered in the IMPACT trial was patients’ acceptance to the CBM intervention. Of the 68 patients enrolled in the study, only 43 completed the six weeks of training, and only 17 provided valid accelerometer data for the week following the rehabilitation programme. This high attrition rate (75%), combined with the already limited enrolment, resulted in a severely underpowered study, with an estimated power of 10% rather than the planned 90%. However, it is worth noting that such high attrition rates are not uncommon in studies evaluating CBM interventions targeting health behaviours such as gambling,^45^ smoking,^46,47^ or alcohol consumption.^48^ Attrition rates reported in these studies range from 43%^48^ to 90.1%.^45^ For instance, Snippe et al.^45^ discontinued their study due to a 90.1% attrition rate after three years of data collection. Importantly, many of these studies were conducted online and without supervision, which likely compounded the challenge of maintaining participants’ motivation.^45^ By contrast, in our study, the CBM tasks were supervised by research assistants, suggesting that even direct oversight may not fully mitigate challenges to engagement.

In the broader context of cardiac rehabilitation programmes, attrition is also a pervasive challenge. Systematic reviews report dropout rates ranging from 12% to 82% across international cardiac rehabilitation programmes.^49–52^ with substantial heterogeneity attributable to differences in programme design, healthcare system factors, and patient characteristics. Higher attrition rates are consistently observed among older patients, women, individuals with multiple comorbidities, and those from underserved racial, social and ethnic minority groups.^49–52^ With an observed attrition rate of 75%, our study falls at the higher end of both CBM intervention studies (43–90%) and cardiac rehabilitation programmes (12–82%). This suggests that the challenges we encountered may be due to the combination of two context-specific difficulties: implementing a repetitive, cognitively demanding digital intervention in a rehabilitation setting, where sustained engagement is already challenging.

Framing our findings within this broader literature highlights that acceptance and adherence are key implementation issues for both digital behaviour-change interventions and standard cardiac rehabilitation services.

One possible explanation for the low acceptance observed in the trial lies in cognitive and emotional indicators. Patients reported difficulty understanding how completing the task could influence their PA behaviour. Additionally, patients expressed that the task felt long and boring and that the images were depressing. The combination of these cognitive (i.e., perceived effectiveness, task irrelevance) and emotional (i.e., boredom) barriers highlights the importance of designing CBM tasks that are perceived as both engaging and relevant.^34,53^ Regarding perceived uselessness, novel digital interventions, such as ours, may be perceived as ineffective in promoting behaviour change – in the absence of direct experiences with the targeted behaviour (e.g., practicing PA in a park, under the supervision of a trained coach).

This low perceived usefulness could stem from a general underestimation of the impact of conditioning procedures on behaviour, coupled with an overestimation of individuals’ ability to consciously self-regulate and control their actions.^9,54^ Addressing this issue is particularly challenging because one of the core assumptions of digital interventions is their ability to influence behaviour without participants needed to fully understand or be aware of the underlying mechanisms.^9^ However, this assumption has been challenged by Van Dessel et al.,^55^ who suggest that the effectiveness of such interventions may also rely on participants’ awareness of the intervention’s goals, thereby engaging more reflective processes.

To increase engagement to training sessions, it is critical to ensure that the subjective value of the task outweighs its associated costs by addressing concerns of boredom and perceived lack of relevance (Table 1). One promising approach is to incorporate meaningful goals and consequences into the intervention.^55^ For instance, in the Antecedents Behaviours Consequences (ABC) training task, participants move an avatar away from unhealthy choices (e.g., fatty foods, cigarettes, alcohol) and towards healthier alternatives, while receiving positive feedback on personally relevant goals and outcomes, such as weight loss or improved health.^56^ Recent applications of ABC training in the context of addiction^57^ and PA^29^ have shown promise. Thus, this type of task may reduce the cognitive and emotional indicators of low acceptance of the IMPACT trial.

Perceived boredom can be attributed to at least two key factors: the repetitive nature of the task and the lack of intrinsically engaging content. While reducing the number of repetitions is challenging – especially since we had already reduced the duration of the CMB while ensuring an adequate dose of intervention – alternative strategies could be explored. For example, the Heartphone mobile application, developed by Conroy and Kim,^58^ exemplifies such a strategy. This application pairs images of PA with positive affective stimuli whenever users interact with their smartphone, fostering an automatic association between PA and positive feelings. This method offers frequent exposure to intervention content without placing additional burden on participants.

To further mitigate boredom, it is important to recognise that highly repetitive tasks can be disengaging, even over short periods of time.^59^ A key lesson learned from this trial is that expecting patients’ adherence to a task that does not provide immediate rewarding value is unrealistic. One promising solution is to incorporate more intrinsically motivating tasks.

For example, the ABC training task can include animations and “vitality gauges” that can increase or decrease depending on the action (e.g., approaching or avoiding PA).^29^ The gamified nature of this task enhances its intrinsic value compared to traditional approach-avoidance tasks.^60,61^ Additionally, emerging technologies such as virtual reality offer opportunities to create more engaging and intrinsically motivating CBM tasks.^62,63^ Consequently, exploring tasks that naturally resonate with participants and leverage automatic responses to exercise-related stimuli represents a valuable avenue for future research.

However, it is also crucial to ensure that participants, regardless of their age, socio-economic background, or health conditions, possess adequate skills to engage with digital interventions.^64^ Addressing this challenge requires a systematic and participatory approach, as outlined in the Medical Research Council (MRC) Framework for developing and evaluating complex interventions.^65^ According to this framework, successful interventions must be developed through iterative phases that prioritise not only efficacy, but also contextual factors (e.g. alignment with participants’ routines, cultural norms and resource availability), stakeholder engagement and adaptability. Therefore, complex interventions, particularly those involving digital tools, must be developed with a deep understanding of the contextual factors influencing their acceptance and feasibility. To address this challenge, the co-production of the protocol by patients, health professionals, researchers, and PPI members, alongside conducting pilot studies prior to full implementation, can help ensure that interventions are accessible, engaging, and effective for the target population.^65–67^ For instance, qualitative research conducted during the feasibility phase may reveal disparities in digital literacy, varying levels of technological proficiency, or cultural preferences that could impact engagement. Adopting the MRC approach, which includes development, feasibility testing, evaluation and implementation, enables future research to systematically integrate stakeholder feedback and contextual adaptations. This approach ensures that digital interventions are technically accessible, culturally appropriate, and feasible for all participants, thereby enhancing their acceptance and impact.

Implementing these recommendations would improve the quality of research, but careful planning and resource allocation are required. Therefore, researchers should explicitly outline these steps in grant proposals and justify the need for additional time, funding and personnel to conduct qualitative research, pilot studies and stakeholder engagement.

Although these measures would extend trial timelines and increase costs, they are essential for maximising the rigour, relevance and impact of complex interventions. Grant institutions should invest in these processes, recognising that they will yield higher-quality, more informative research and ultimately deliver greater value for public and scientific benefit.

### Strengths and Limitations

This retrospective study has several important limitations. First, the data analysed were not collected systematically with the purpose of evaluating acceptance. Rather, they emerged from informal observations of the implementation process relayed by research assistants during the original trial. The absence of standardised procedures for gathering and documenting this feedback increases the risk of incomplete or selective reporting, which may compromise the robustness of our conclusions. Second, our analysis is limited by the small sample size and low engagement rates, which restrict our ability to draw definitive conclusions and generalisability about the reasons for CBM intervention failure. Despite these limitations, the informal feedback provided by research assistants serves as a valuable pillar of this study. These observations provide valuable insights into the barriers to implementation that could inform the design of future CBM interventions in cardiac rehabilitation. Building on these findings, we recommend that future trials incorporate validated acceptance frameworks, systematic feedback mechanisms and qualitative research prior to implementation, in order to better anticipate and mitigate potential barriers and prevent implementation failures.

## Conclusion

This study highlights both the challenges and opportunities in developing and implementing CBM-based PA interventions in cardiac rehabilitation. The barriers identified, including perceived task irrelevance, monotony, implementation burden, and limited family support, provide a roadmap for future intervention. Effective PA promotion requires approaches that are not only evidence-based but also acceptable, engaging, feasible, and seamlessly integrated into both patients’ lives and health care routines. By openly sharing these implementation lessons, we aim to accelerate the development of more effective digital interventions that can maximise the significant PA benefits of enhancing the health and quality of life of cardiovascular patients.

## Supporting information

Descriptive characteristics of the sample are presented in Supplementary Material 1.

## Data Availability

Deidentified data, data management, analysis codes and research materials have been made publicly available on Zenodo

https://doi.org/10.5281/zenodo.17292448

## Availability of Data and Materials

Deidentified data, data management, analysis codes and research materials have been made publicly available on Zenodo.^68^

## Acknowledgements

The authors are grateful to Arbnor Hasani, Auriane Emeline Micheli, Zoë Alicia Piazza of the University of Geneva, and Anaïs Quossi of the University of Lyon for their implication on data collection. The authors are grateful to all study participants who volunteered, as well as the medical staff at Geneva University Hospitals for their assistance.

## Authors contributions

LF: Conceptualisation, investigation, formal analysis, writing-original draft, writing-review and editing; SM: Methodology-creation of the tasks, writing-review and editing; PM; Resources, writing-review and editing; AF: Writing-review and editing; SC: writing-review and editing; DS: writing-review and editing; MF: writing-review and editing; RWW: writing-review and editing; AF: Conceptualisation, writing-review and editing; CL: Resources, writing-review and editing; PS: Supervision, writing-review and editing; BC: Conceptualisation, supervision, writing-original draft, writing-review and editing, guarantor.

## Footnotes

a Of the planned sample size of 352 patients, we reached 19% (n = 68) of the target enrolment, 12% (n = 43) for engagement in the CBM training, and a mere 5% (n = 17) for PA data in the week following discharge from the rehabilitation programme

