## Supplementary material for "Anatomy of a Failure: A Retrospective Evaluation of a Cognitive Bias Modification Intervention to Promote Physical Activity in Cardiac Rehabilitation": Descriptive characteristics of the sample are presented in Supplementary Material 1.

**Supplementary Materials**

**Supplementary Material 1.** Descriptive Table of the Outpatients at Baseline

**Supplementary Material 2.** Comparison Between the Intervention and the Placebo Groups

**Supplementary Materials 3.** Post-hoc power analysis

**Supplementary Materials 4.** Cognitive Indicator

**Supplementary Material 5.** Description of the Outpatient Cardiac Rehabilitation Programme

**Supplementary Material 1.** Descriptive Table of the Outpatients at Baseline

**Table S1.** Descriptive Characteristics of the Patients at the Beginning of the Cardiac Rehabilitation Programme

| N = 68 | M (SD) |
| --- | --- |
| Demographics and anthropometrics |  |
| Sex (*n*, %) |  |
| Female | 9 (13.24%) |
| Male | 59 (86.74%) |
| Age (years, *n*, %) |  |
| <50 | 14 (20.90%) |
| 50–59 | 25 (37.31%) |
| 60–69 | 17 (25.37%) |
| 70–79 | 10 (14.93%) |
| >80 | 1 (1.49%) |
| Body mass index (Kg/m^2^; *n*, %) |  |
| Underweight <18.5 | 0 (0%) |
| Normal 18.5–<25 | 19 (27.79%) |
| Overweight 25–<30 | 30 (45.45%) |
| Obese ≥ 30 | 17 (25.76%) |
| Usual level of PA (*n*, %) |  |
| Physically inactive | 18 (26.47%) |
| Some PA | 28 (41.18%) |
| Regular PA | 20 (29.41%) |
| High training | 2 (2.94%) |
| Emotional health-related variables |  |
| Anxiety [1–5] | 1.79 (0.90) |
| Depressive symptoms [1–5] | 1.62 (0.76) |
| Fatigue [1–5] | 2.55 (1.06) |
| Pain intensity [1–5] | 2.10 (2.29) |
| Additional health-related variables |  |
| Number of ADL (*n*, %) |  |
| No ADL | 59 (86.76%) |
| ≥1 ADL | 9 (13.24%) |
| Number of IADL (*n*, %) |  |
| No IADL | 49 (72.13%) |
| ≥1 IADL | 18 (26.87%) |
| Mobility [1–5] | 3.70 (0.54) |
| Global physical health [1–5] | 2.88 (0.89) |
| Global mental health [1–5] | 3.77 (0.98) |
| Life satisfaction [1–7] | 5.68 (1.26) |
|  | *Continued* |

**Table S1.** (*Continued*)

| N = 68 | M (SD) |
| --- | --- |
| LVEF (%) | 52.87 (10.49) |
| MAP (Watt) | 141.08 (45.08) |
| Abdominal circumference (cm) | 99.31 (12.02) |
| HbA1c (%) | 5.69 (0.74) |
| Triglycerides (mmol/L) | 1.59 (1.08) |
| LDL (mmol/L) | 2.94 (1.25) |
| Systolic blood pressure (mmHg) | 126.10 (14.64) |
| Diastolic blood pressure (mmHg) | 72.49 (8.29) |
| Cigarette (pack/year) | 10.82 (14.85) |
| Comorbidity (*N* = 67, *n*, %) |  |
| 0 | 9 (13.43%) |
| 1 | 18 (26.87%) |
| ≥ 2 | 40 (59.70%) |
| Indication for enrollment in the outpatient CR (*N* = 67, *n*; %) |  |
| Acute coronary syndrome NSTEMI | 27 (40.30%) |
| Acute coronary syndrome STEMI | 18 (26.87%) |
| Acute coronary syndrome UA | 4 (5.97%) |
| Coronary artery bypass graft surgery | 7 (10.45%) |
| Coronary artery disease without ACS or surgery | 8 (11.94%) |
| Heart failure | 2 (2.99%) |
| Valvular surgery | 1 (1.49%) |
| Motivational variables towards PA |  |
| Perceived capabilities [1–5] | 4.21 (0.97) |
| Intention [1–10] | 5.51 (1.34) |
| Instrumental attitudes [1–10] | 9.09 (1.42) |
| Affective attitudes [1–10] | 8.16 (2.06) |
| Approach toward PA | -34.22 (109.97) |
| Approach toward SB | 8.98 (102.38) |

Abbreviations: M, mean; SD, standard deviation; PA, physical activity; ADL, limitations in activities of daily living; IADL, limitations in instrumental activities of daily living; CR; cardiac rehabilitation; NSTEMI, non-ST elevation myocardial infarction; STEMI, ST elevation myocardial infarction; UA, unstable angina; ACS, acute coronary syndrome; LVEF, left ventricular ejection fraction; MAP, maximal aerobic power; HbA1c, fasting glycated hemoglobin; LDL, low-density lipoprotein-cholesterol; SB, sedentary behaviors. Usual level of PA was assessed with the Saltin-Grimby PA Level Scale. The LVEF is considered to fall within the normal range when it is between 50% and 70% [92]. A waist circumference greater than 95 cm for men and 80 cm for women is associated with an increased risk of all-cause mortality [93]. HbA1c levels are classified as normal, or within the non-diabetic range below 5.7% [94]. Triglycerides levels are considered normal if they are below 1.7 mmol/L [95]. LDL cholesterol levels are desirable if they are below 1.8 mmol/L for individuals with coronary artery disease or other forms of atherosclerosis, and below 2.6 mmol/L for healthy individuals [96]. Systolic blood pressure is considered high if it exceeds 129 mmHg, and diastolic blood pressure is considered high if it exceeds 79 mmHg [97]. A positive score in *Approach toward PA* suggests a tendency to approach PA, while a negative score suggests a tendency to avoid PA. A positive score in *Approach toward SB* suggests a tendency to approach PA, while a negative score suggests a tendency to avoid PA. All information on variables assessment methods can be found in Fessler et al. (2024).

Fessler, L., Tessitore, E., Craviari, C. *et al.* Motivational and emotional correlates of physical activity and sedentary behavior after cardiac rehabilitation: an observational study. *BMC Sports Sci Med Rehabil* **16**, 209 (2024). https://doi.org/10.1186/s13102-024-00997-0

**Supplementary Material 2.** Comparison Between the Intervention and the Placebo Groups

**Table S2.** Baseline Characteristics of Participants with Complete Post-Intervention Physical Activity Data (N = 17)

| ***N* = 17** | **Intervention**  ***N* = 10** | **Missing data** | **Placebo**  ***N* = 7** | **Missing data** |  |
| --- | --- | --- | --- | --- | --- |
|  | *M (SD)* | *N* | *M (SD)* | *N* | *P* value* |
| Demographics and anthropometrics |  |  |  |  |  |
| Sex (*n*, %) |  |  |  | 1 |  |
| Female | 4 (40%) | 0 | 1 (14%) | 0 |  |
| Male | 6 (60%) | 0 | 6 (86%) | 0 | .338 |
| Age (years, *n*, %) |  |  |  |  | .815 |
| <50 | 1 (10%) | 0 | 1 (14%) | 0 |  |
| 50–59 | 4 (40%) | 0 | 2 (29%) | 0 |  |
| 60–69 | 4 (40 %) | 0 | 2 (29%) | 0 |  |
| 70–79 | 4 (40%) | 0 | 2 (29%) | 0 |  |
| >80 | 0 (0%) | 0 | 0 (0%) | 0 |  |
| Body mass index (Kg/m^2^; *n*, %) |  |  |  |  | .079 |
| Underweight <18.5 | 0 (0%) | 0 | 0 (0%) | 0 |  |
| Normal 18.5–<25 | 5 (50%) | 0 | 5 (71%) | 0 |  |
| Overweight 25–<30 | 4 (40%) | 0 | 2 (29%) | 0 |  |
| Obese ≥ 30 | 1 (10%) | 0 | 0 (0%) | 0 |  |
| Usual level of PA (*n*, %) |  |  |  |  | .50 |
| Physically inactive | 2 (20%) | 0 | 0 (0%) | 0 |  |
| Some PA | 4 (40%) | 0 | 5 (71%) | 0 |  |
| Regular PA | 4 (40%) | 0 | 2 (29%) | 0 |  |
| High training | 0 (0%) | 0 | 0 (0%) | 0 |  |
| Emotional health-related variables |  |  |  |  |  |
| Anxiety [1–5] | 1.65 (0.91) | 0 | 1.64 (0.85) | 0 | .987 |
| Depressive symptoms [1–5] | 1.68 (0.86) | 0 | 1.39 (0.48) | 0 | .445 |
| Fatigue [1–5] | 2.27 (0.86) | 0 | 2.10 (1.05) | 0 | .716 |
| Pain intensity [1–5] | 1.90 (1.79) | 0 | 0.86 (1.57) | 0 | .234 |
| Additional health-related variables |  |  |  |  |  |
| Number of ADL (*n*, %) |  |  |  |  | .412 |
| No ADL | 10 (100%) | 0 | 6 (86%) | 0 |  |
| ≥1 ADL | 0 (0%) | 0 | 1 (14%) | 0 |  |
| Number of IADL (*n*, %) |  |  |  |  | .644 |
| No IADL | 7 (70%) |  | 4 (57%) |  |  |
| ≥1 IADL | 3 (30%) | 0 | 3 (43%) | 0 |  |
| Mobility [1–5] | 3.80 (0.42) | 0 | 4.00 (0.00) | 0 | .143 |
| Global physical health [1–5] | 2.50 (0.71) | 0 | 3.71 (1.11) | 0 | .015 |
| Global mental health  [1–5] | 3.60 (1.08) | 0 | 4.29 (0.76) | 0 | .168 |
| Life satisfaction [1–7] | 2.30 (0.82) | 0 | 1.71 (0.49) | 0 | .114 |
|  |  |  |  |  | *Continued* |

**Table S2.** (*Continued*)

| ***N* = 17** | **Intervention**  ***N* = 10** | **Missing data** | **Placebo**  ***N* = 7** | **Missing data** |  |
| --- | --- | --- | --- | --- | --- |
|  | *M (SD)* | *N* | *M (SD)* | *N* | *P* value* |
| LVEF (%) | 52.50 (12.10) | 2 | 55.00 (9.75) | 1 | .686 |
| MAP (Watt) | 124.67 (63.74) | 1 | 159.86 (45.85) | 0 | .239 |
| Abdominal circumference (cm) | 90.50 (8.85) | 0 | 93.00 (9.31) | 0 | .583 |
| HbA1c (%) | 5.69 (1.01) | 3 | 5.30 (0.34) |  | .358 |
| Triglycerides (mmol/L) | 1.27 (0.45) | 1 | 1.55(0.81) | 0 | .389 |
| LDL (mmol/L) | 2.99 (1.17) | 0 | 3.65 (0.55) | 0 | .187 |
| Systolic blood pressure (mmHg) | 123.40 (11.84) | 0 | 120.86 (15.31) | 0 | .704 |
| Diastolic blood pressure (mmHg) | 72.80 (7.90) | 0 | 67.29 (8.79) | 0 | .196 |
| Cigarette (pack/year) | 4.50 (7.62) | 0 | 5.14 (8.86) | 0 | .875 |
| Comorbidity (*n*, %) |  |  |  | 0 |  |
| 0 | 2 (20%) | 0 | 0 (0%) | 0 | .647 |
| 1 | 4 (40%) | 0 | 4 (57%) | 0 |  |
| ≥ 2 | 4 (40%) | 0 | 3 (43%) | 0 |  |
| Indication for enrollment in the outpatient CR (*n*; %) |  |  |  |  | 1.000 |
| Acute coronary syndrome NSTEMI | 4 (40%) | 0 | 3 (43%) | 0 |  |
| Acute coronary syndrome STEMI | 2 (20%) | 0 | 2 (28.5%) | 0 |  |
| Acute coronary syndrome UA | 1 (10%) | 0 | 2 (28.5%) | 0 |  |
| Coronary artery bypass graft surgery | 0 (0%) | 0 | 0 (0%) | 0 |  |
| Coronary artery disease without ACS or surgery | 1 (10%) | 0 | 0 (0%) | 0 |  |
| Heart failure | 0 (0%) | 0 | 0 (0%) | 0 |  |
| Valvular surgery | 2 (20%) | 0 | 0 (0%) | 0 |  |
| Motivational variables towards PA |  |  |  |  |  |
| Perceived capabilities [1–5] | 4.40 (0.70) | 0 | 4.71 (0.49) | 0 | .340 |
| Intention [1–10] | 5.65 (1.13) | 0 | 4.86 (2.12) | 0 | .331 |
| Instrumental attitudes [1–10] | 9.10 (1.73) | 0 | 9.71 (0.76) | 0 | .395 |
| Affective attitudes  [1–10] | 8.90 (1.73) | 0 | 9.00 (1.16) | 0 | .896 |
| Approach toward PA | -20.20 (87.36) | 3 | 26.41 (112.71) | 0 | .404 |
| Approach toward SB | -37.836 (107.02) | 3 | 41.67 (85.07) | 0 | .150 |

Abbreviations: M, mean; SD, standard deviation; PA, physical activity; ADL, limitations in activities of daily living; IADL, limitations in instrumental activities of daily living; CR; cardiac rehabilitation; NSTEMI, non-ST elevation myocardial infarction; STEMI, ST elevation myocardial infarction; UA, unstable angina; ACS, acute coronary syndrome; LVEF, left ventricular ejection fraction; MAP, maximal aerobic power; HbA1c, fasting glycated hemoglobin; LDL, low-density lipoprotein-cholesterol; SB, sedentary behaviors. Usual level of PA was assessed with the Saltin-Grimby PA Level Scale. *P-values from independent samples t-tests (numerical variables) or Fisher’s exact test (categorical variables). These analyses are severely underpowered (total n = 17) and should be interpreted only as descriptive information. The study experienced 75% attrition and substantial implementation challenges. These results do not provide meaningful evidence regarding intervention effectiveness.

**Table S3.** Accelerometer-Measured Physical Activity One Week After Discharge from the Programme: Between-Group Comparisons (N = 17)

| **N = 17** | **Intervention**  **N = 10** | **Missing data** | **Placebo**  **N = 7** | **Missing data** |  |
| --- | --- | --- | --- | --- | --- |
|  | M (SD) | N | M (SD) | N | *P* value* |
| Daily MVPA (min/day) | 60.43 (43.56) | 0 | 59.91 (19.52) | 0 | .977 |
| Daily sedentary behaviour (min/day) | 554.16 (72.03) | 0 | 540.89 (35.59) | 0 | .661 |

Abbreviations: M, mean; MVPA: moderate-to-vigorous physical activity. * P-values from independent samples t-tests

**Supplementary Materials 3.** Post-hoc power analysis

**Figure S1**
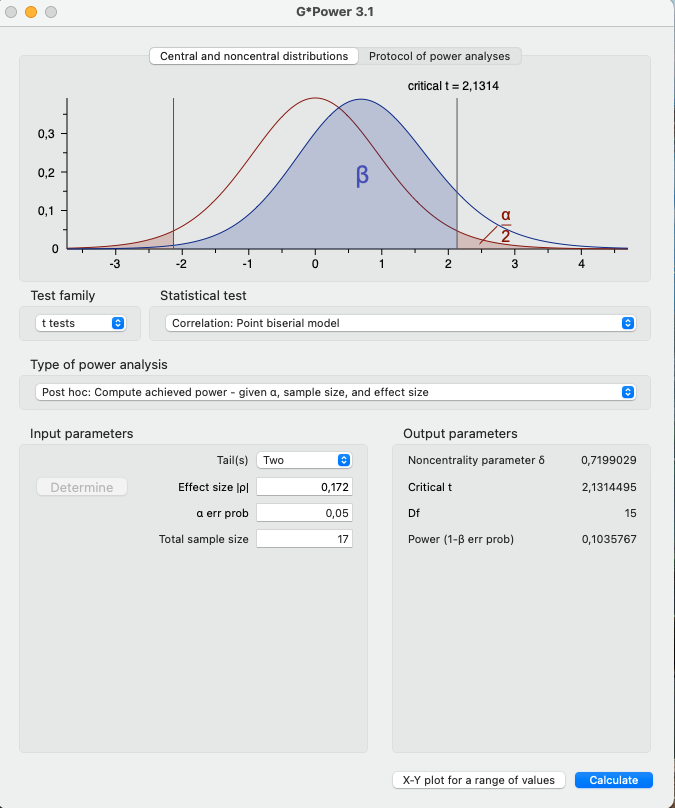
**.** Post-Hoc Analysis

**Supplementary Materials 4.** Cognitive Indicator

**Table S4.** Cognitive Indicator

| **Outcome** | **Mean (sd)** | **Min-Max** |
| --- | --- | --- |
| Perceived effectiveness of the intervention [1–5] | 3.09 (0.91) | 1–5 |

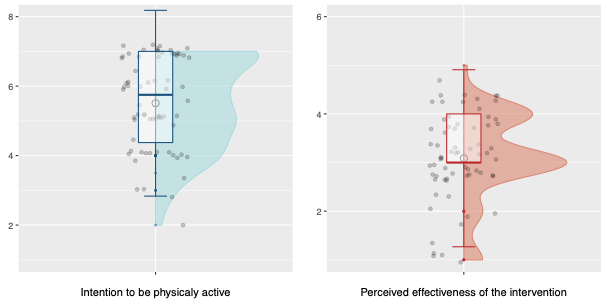
**Figure S2.** Perceived Effectiveness of the CBM Intervention

**Supplementary Material 5.** Description of the Outpatient Cardiac Rehabilitation Programme

The outpatient cardiac rehabilitation (CR) programme adheres to the guidelines established by the Swiss Working Group for Cardiovascular Prevention, Rehabilitation, and Sports Cardiology (SCPRS), as outlined on www.SCPRS.ch. These guidelines are based on the core components of CR, which were recently updated by the European Association of Preventive Cardiology.^1^ The ambulatory or outpatient CR programme takes place over a period of 6 weeks, with sessions scheduled every morning of the 5 weekdays. The programme comprises overall 78 training sessions of 45 minutes that emphasise endurance, strength, coordination/balance, outdoor activities, and relaxation techniques. In addition, there are also 12 educational sessions that provide information and material on disease comprehension, lifestyle modifications, medication information, dietary guidance, smoking cessation support, diabetic counselling, stress management, and psychological assessment when required. The outpatient CR programme is not recommended for individuals dealing with complicated clinical or psychosocial situations, such as advanced age, reduced physical ability, impaired left ventricular function, comorbidities, or stressful home environments who are instead directed to an inpatient program.
